# Social prescribing for patients with severe mental illnesses: an English primary care longitudinal observational study

**DOI:** 10.64898/2026.10.01.26364429

**Authors:** Alexandra Burton, Naomi Launders, Amy E Taylor, Alvin Richards-Belle, David Osborn, Stephanie Tierney, Daisy Fancourt, Feifei Bu

## Abstract

**Background:** People diagnosed with severe mental illnesses (SMI) including bipolar disorder, schizophrenia and psychosis are disproportionately exposed to adverse social determinants of health, contributing to premature morbidity and mortality. Social prescribing aims to address social risk factors for poor health by linking patients in primary care to community activities. However, whether people with SMI access social prescribing through primary care remains unexplored.

**Aims:** To describe prevalence of, and sociodemographic characteristics associated with, social prescribing consultations in patients with SMI in primary care.

**Methods:** Using the UK Clinical Practice Research Datalink, we identified patients aged 18+ with a diagnosis of SMI between 2004-2019. We calculated the percentage of patients with SMI who received a social prescribing consultation between 2019-2023 and used logistic regression to examine associations between sociodemographic, health-related factors and social prescribing consultations and refusals. We used fixed effects analysis to determine whether a new diagnosis of SMI made between 2019-2023 was associated with an increased odds of receiving a social prescribing consultation.

**Results:** Among 112,334 patients with SMI, 0.8% received a social prescribing consultation in 2019 increasing to 9.5% in 2023. Between 2020-2023, older age (OR=1.18-2.34), being female (OR=1.16-1.31) and living in more deprived areas (OR=1.14-1.49) were associated with higher odds of receiving a social prescribing consultation amongst patients with SMI. Across all years, patients with SMI and depression (OR=1.32-1.92), diabetes (OR=1.19-1.35) or obesity (OR=1.17-1.54) also had increased odds. In 140,514 patients with time-varying SMI status, a new diagnosis of SMI was associated with higher odds of receiving a social prescribing consultation (OR=1.39, 95% CI 1.26-1.53).

**Conclusion:** Since 2019, social prescribing consultations among patients with SMI in primary care have increased and are reaching those in areas of higher deprivation and with greater health needs. Strategies to remove barriers to consultations among younger adults and males are still needed.

## Introduction

People living with severe mental illnesses (SMI) including schizophrenia, bipolar disorder and other psychotic illnesses, experience stark health inequalities, characterised by high rates of physical health problems(1) that lead to an increased risk of premature mortality compared to the general population(2–4). Associations between SMI diagnoses and physical multimorbidity have been identified in the context of intersecting vulnerabilities such as deprivation and ethnicity(5); increased risk of cardiovascular disease risk factors such as smoking(6, 7), obesity, hypertension and diabetes(7); increased rates of depression(8); and high rates of social isolation and exclusion(9), all of which further exacerbate the risk of morbidity and early mortality(10). This cumulative impact of mental health, physical health, social and economic disadvantage alongside barriers to accessing preventative healthcare services(11, 12) underscores a pressing need for holistic models of support and care for people with SMI. Social prescribing is one such model that has been implemented across UK primary care services as a policy response aimed at helping to address specific social determinants of health and tackling health inequalities(13). It is recommended for people living with long term health conditions, depression and anxiety and those with complex social needs(14). Social prescribing typically involves a link worker working with patients to determine what matters to them and connecting them to non-clinical community-based resources such as welfare, peer and volunteering support, arts, physical activities and nature-based groups. While effective clinical programmes exist within the UK National Health Service (NHS) for supporting smoking and alcohol cessation, other target behaviours for disease prevention which disproportionately affect people with SMI are increasingly being supported through social prescribing pathways. Evidence from a scoping review found that diabetes was the most commonly cited health condition in people with long term health conditions referred to social prescribing(15). Social prescribing has also been identified as a promising intervention for improving health behaviours associated with diabetes, obesity and hypertension(16) as well as for alleviating depression(17) and reducing loneliness(18). As a result, social prescribing has been proposed as an approach to improving the physical health and wellbeing of people with SMI(9, 19, 20), which could be particularly important during the early stages of their diagnosis to help prevent modifiable risk factors for poor health from developing and becoming entrenched.

However, there remains uncertainty over whether social prescribing is equitable in its reach. People with SMI have reported barriers to accessing social prescribing including experiences of stigma and social anxiety(21, 22), as well as practical barriers such as financial constraints for attending end-point community activities(9, 22). General practitioners highlight practical barriers to referral and engagement in social prescribing activities for patients with mental health problems more broadly, including limited consultation appointment times to fully explore patient needs and variable local provision of community programmes(23). At a system level, evidence suggests that the rollout of link workers does not match population need, with under-provision in the most deprived and ethnically diverse areas where SMI diagnoses are likely to be higher and support needs greater(24, 25). Qualitative work with social prescribing link workers has also identified a lack of training and confidence in working with people with complex and severe mental health problems which could lead to premature referral back to clinical services and patient disengagement with social prescribing(25, 26). Against this backdrop, there is a need for population-level evidence on whether people with SMI are being offered social prescribing within primary care to clarify the prevalence of social prescribing activity for this population. This study therefore aims to i) describe the proportion of people diagnosed with SMI recorded as having a social prescribing consultation in their primary care medical record, ii) explore sociodemographic and health factors associated with social prescribing consultations and refusals among people with SMI, and iii) determine whether receiving a new diagnosis of SMI is associated with social prescribing consultation.

## Methods

### Data

Data were from the Clinical Practice Research Datalink (CPRD); a research data service that routinely collects anonymised patient data from over 2000 GP practices across the UK. This study focused on data from CPRD Aurum. At the time of data extraction for this study, CPRD Aurum contained data on demographics, diagnoses and symptoms, prescriptions, tests and referrals from 47.8 million patients over more than 35 years since 1989, including 16.2 million currently registered patients (June 2024 version). It covered 19.9% of general practices in England and has been shown to be representative of the English population in terms of geographic area, deprivation, urbanicity, age and sex (27, 28).

### Study cohort

The study cohort was defined as patients aged 18 or over on the SMI Quality and Outcomes Framework (QOF) register with a SNOMED CT code for SMI (schizophrenia, bipolar disorder or other non-organic psychotic illness) recorded between the period 1 January 2004 to 31 December 2023. In total, this provided an initial sample of 227,259 patients from 1,783 GP Practices. The study follow-up period was restricted to 2019-2023, starting from the year in which social prescribing was formally rolled out within the NHS(29) and excluding incomplete data for the year 2024. To determine the proportion of patients with SMI with a social prescribing consultation and the sociodemographic and health factors associated with social prescribing consultations and refusals, we restricted the sample to adult patients with an SMI diagnosis before 2019 who were alive and still registered at the end of the study follow-up period (31 December 2023) and who had at least one consultation recorded in their medical record in any year between 2019-2023. This provided an analytical sample of 112,334 patients (cohort 1).

To determine whether having a more recent diagnosis of SMI was associated with receiving a social prescribing consultation, we extended the sample to also include patients with an SMI diagnosis first recorded between the period 2019-2023. After excluding those who had died up to 31^st^ December 2023 and those without a GP consultation within the study period (2019-2023), we had a sample of 140,514 patients (cohort 2) with time-varying SMI status which included patients with an SMI diagnosis prior to 2019 and a comparator group of patients with no previous record of SMI who went on to receive an SMI diagnosis between 2019-2023 (Supplementary Materials 1, Figure S1).

### Outcomes

Consultations related to social prescribing were identified using the code lists presented in table S1, which included any code related to social prescribing consultation(30). These codes ranged from ‘referral to social prescribing service’, ‘social prescribing offered’, to ‘signposting to social prescribing service’. Consultation was defined in a broad sense, including contacts with either clinical or non-clinical staff members. Social prescribing service refusal was defined as having a code of ‘social prescribing declined’ recorded in the patient’s medical record.

### Covariates

We included the following sociodemographic factors as covariates: age groups (in 2018) derived from year of birth and categorised as ages 18-29, 30-39, 40-49, 50-59, 60-69, 70-79 or 80+, sex (male, female), ethnicity categorised as white or ethnic minority; ethnicity was initially coded into Asian, black, mixed/other and white, which was recoded into binary codes because of small numbers in each category, area deprivation in quintiles measured by the 2019 index of multiple deprivation (IMD) based on patient postcode, and area urbanicity (rural, urban) based on the Rural Urban Classification from the 2011 census. Health-related factors identified in the literature as being amenable to change in the general population through social prescribing(15–17) and that are also experienced disproportionately among people with SMI(7, 8) included obesity, defined as having a SNOMED code for the Obesity QOF register or a Body Mass Index (BMI) greater or equal to 30, compared to under/normal weight (<25) and overweight (25-29.9), diabetes, hypertension or depression diagnoses, which were identified using SNOMED codes for the relevant QOF registers (for code lists see Supplementary Materials 2). These health-related covariates were treated as baseline measures indicating whether patients ever had each condition before 2019.

### Statistical Analysis

To assess the reach of social prescribing, we calculated the percentage of patients with a social prescribing consultation among all patients diagnosed with SMI with a GP consultation by calendar year between 2019 and 2023. We then examined whether sociodemographic and health-related factors were associated with social prescribing consultation and social prescribing service refusal using multilevel logistic regression models, accounting for the nested data structure (patients within primary care practices). The intraclass correlation coefficient (ICC) and median odds ratio (MOR), estimated from unconditional models, were used to quantify between-practice variance in the outcome. Analyses were stratified by year.

To determine whether a new SMI diagnosis was associated with social prescribing consultation we used fixed effects analysis, treating SMI diagnosis and social prescribing consultations as time-varying variables where patients may have repeated GP visits across years. Fixed effects analysis uses only within-person variation, automatically controlling for all observed and unobserved time-invariant covariates, estimating an average within-person association between a time-varying predictor (diagnosis of SMI during the study period) and the outcome (social prescribing consultation). We included year dummies to control for potential time trends and used cluster-robust standard errors to account for residual correlations within practices.

Missing data were minimal for most sociodemographic covariates except for area deprivation and urbanicity (18.5% in analytical cohort one and 18.9% in cohort two) that were obtained via postcode linkage (Supplementary Materials 1, Table S2). To deal with missing data, we carried out multiple imputation (MI) with chained equations (MI=20) including all variables in the analyses. All analyses were conducted in Stata V18.

### Ethics and consent

The authors assert that all procedures contributing to this work comply with the ethical standards of the relevant national and institutional committees on human experimentation and with the Helsinki Declaration of 1975, as revised in 2013. Ethical approval for this study was obtained from the Independent Scientific Advisory Committee of CPRD (protocol number 24_004142). All CPRD data are anonymised and therefore participant consent is not required.

## Results

The demographic and health characteristics of the study cohorts are presented in Table 1. In analytical cohort 1, 24.7% of patients with SMI were aged 60 or above, 52.6% were female, 23.1% were from ethnic minority backgrounds and 10.1% lived in rural areas. Patients from the top two most deprived quintiles were overrepresented. Regarding health characteristics, 37.1% of the cohort had obesity, 13.2% had a SNOMED code for the diabetes register, 17.6% had a SNOMED code for the hypertension register, and 45.7% had a SNOMED code for the depression register. Analytical cohort 2 had a similar profile.

As shown in Figure 1, 0.8% of patients with SMI received a social prescribing consultation in 2019, which steadily increased to 9.5% in 2023. The ICC decreased from 0.5 in 2019 to 0.2 in 2023, and MOR from 5.7 to 2.5, suggesting that between-practice variation in social prescribing consultations decreased over time.

Figure 2 shows the results from multilevel logistic regression models on social prescribing consultations (see also Supplementary Materials 1, Table S3). In the 2019 model, there was some, but limited evidence that sociodemographic factors were associated with social prescribing consultation for patients with SMI. However, between 2020 and 2023, patients with SMI aged 40 or above were consistently more likely to receive a social prescribing consultation than patients with SMI under 30 years old (OR=1.18-2.34). Females had 16-31% higher odds of receiving a social prescribing consultation compared to males and patients from more deprived areas (third to fifth quintiles) had 14-49% higher odds of receiving a social prescribing consultation compared to patients in the least deprived areas (first quintile). There was no evidence that ethnicity or area urbanicity were related to social prescribing consultations. Regarding specific health factors, higher odds of having a social prescribing consultation was also found across all years amongst people with obesity (OR 1.17-1.54), diabetes (OR 1.19-1.35) and depression (OR 1.32-1.92) and all years apart from 2023 amongst people with hypertension (OR 1.14-1.29).

Overall, 16.1% of people with an SMI diagnosis received a “social prescribing declined” code in 2019, 14.0% in 2020, 9.4% in 2021, 10.1% in 2022 and 10.8% in 2023 (Supplementary Materials 1, Table S4). There was no evidence that any sociodemographic or health factors were associated with service refusal, except for age (Figure 2). Compared to those aged under 30, older age groups, particularly those aged 50 or older, were 2-3 times more likely to decline social prescribing services consistently across all years apart from 2019 where the sample size was small (n=899).

Examining within-person changes across years in social prescribing consultations and SMI diagnosis status, 17.0% of patients had a least one recorded social prescribing consultation, among whom social prescribing was recorded in 28.0% of their observations over time. 14.7% of patients had a new diagnosis of SMI recorded in their medical records between 2019 and 2023 (Supplementary Materials 1, Table S5). Results from the fixed effects models treating SMI as a time-varying variable are presented in Table 2. After accounting for time trends, having a new SMI diagnosis was associated with 39% higher odds of having a social prescribing consultation (OR:1.39, 95%CI: [1.26, 1.53]).

## Discussion

This is the first known study to provide exploratory estimates of social prescribing consultations and refusals among patients diagnosed with SMI in primary care in England. Our findings demonstrate a steady increase in social prescribing among patients with SMI from 0.8% in 2019 (the year that social prescribing was formally launched as an NHS primary care service in England), to 9.5% in 2023. This increase across years is similar to patterns observed in a previous analysis of primary care records in the general population(30) but is comparatively higher than the 2-3% of primary care patients who received a social prescription in 2023(30, 31). Additionally, our analyses focused only on social prescribing consultations within primary care; however evidence suggests that people with SMI may also receive social prescribing referrals via secondary mental health services and upon discharge from inpatient hospital stays(32). Therefore, this figure is likely to be an underestimation.

The findings align with previous research in the general population showing that older age groups and females are more likely to receive social prescribing consultations(30, 33). However, given that the average age of onset for psychosis and schizophrenia is 16-24 years old and 25-44 years old for bipolar disorder(34) and that schizophrenia is more commonly diagnosed in younger men(35), younger people at risk of developing SMI and those early on in their diagnosis are an important target group for social prescribing. Our finding that people receiving a new SMI diagnosis between 2019-2023 had higher odds of receiving a social prescribing consultation than those with a diagnosis received prior to 2019 suggests that GPs may be considering social prescribing as a mechanism for earlier intervention in this group. This could be particularly important for supporting the prevention and worsening of risk factors for physical health problems that lead to increased morbidity and mortality at a younger age than in the general population and that are amenable to change through social prescribing (e.g. loneliness, stress, and low levels of physical activity). Alternatively, this could reflect concerns that social prescribing is being used as a substitute rather than adjunct to specialist mental health care in response to underfunding of healthcare services and inadequate provision of social interventions within statutory mental health care(36). Further work is however needed to understand patterns of healthcare service use among this group and to ensure that people with longer-term diagnoses of SMI are not being overlooked as potential candidates for social prescribing, especially if they do not have frequent contact with their GP.

Across all years, having obesity, diabetes or depression alongside a diagnosis of SMI was associated with higher odds of social prescribing consultation. This is encouraging as it suggests that social prescribing is being used in line with its intended purpose to support those with comorbid physical and mental health conditions(14) and that primary care staff are identifying opportunities to address the wider physical and psychological needs of people with SMI. This is particularly important given the increased risk of morbidity and mortality from cardiovascular disease in this population(3). The analysis also showed that people with SMI living in more deprived areas are more likely to receive a social prescribing consultation. People with SMI are more likely to live in deprived areas and are therefore more likely to experience higher levels of social exclusion, unemployment and financial difficulties(37) suggesting this non-medical support may be appropriately targeting those with the greatest social needs. Given that people with SMI also report barriers to engagement in end-point community activities(22), social prescribing link workers based in resource poor areas may have a particularly key role to play in supporting patients to identify activities of interest to support their health and wellbeing. While there has been an increase in recognition of the role that social prescribing could play in helping to tackle the social and health inequalities experienced by this population, research into its effectiveness is however still in its infancy(21, 38).

Notably, social prescribing refusal rates dropped from 16.1% to 10.8% across the study period, suggesting increased awareness of and acceptability of social prescribing as an intervention to support patients with SMI. This could also indicate improved communication of the potential benefits of social prescribing between clinicians and patients. While older adults were more likely to receive a social prescribing consultation than younger people, the only consistent predictor of refusal was older age, particularly among those ≥50 years. Previous research with older adults has identified mobility issues, difficulties using technology, lack of available transport and anxiety or fear of new environments as barriers to engaging in social prescribing(39), all of which may be compounded by living with an SMI. Older adults with SMI report greater dissatisfaction with healthcare provider communication and shared decision making than older adults without SMI(40) which may influence their likelihood of engaging with new services offered by clinicians. As such, the findings of these age-related inequalities suggest the importance of understanding and overcoming barriers to engagement amongst older adults with SMI.

### Strengths and limitations

A strength of our work is the analysis of a national primary care administrative dataset with an SMI cohort over the five-year period in which social prescribing was formally launched and implemented in England. We compared characteristics of patients with SMI who had a social prescribing consultation recorded in their medical records versus those who had not, which allowed us to understand equity in access among different groups. There are, however, important limitations to the study; physical health related factors and depression diagnoses were defined using QOF registers as we hypothesised that being on the QOF register would increase the likelihood of a patient attending their GP practice for related health checks and therefore also the likelihood of receiving social prescribing. However, the QOF registers represent a subset of all the SNOMED codes available to GPs and therefore the true prevalence of these conditions is likely underestimated. While we were able to explore whether sociodemographic and clinical risk factors for poorer health increased the likelihood of social prescribing consultation for people with SMI, some important risk factors such as loneliness and social isolation remain unexplored due to their limited coding within primary care medical records. Future research should explore the potential for incorporating social health factors into annual health checks for people with SMI to enable a more comprehensive analysis of the impact of loneliness and social isolation on health, and to determine whether social prescribing is reaching one of its key target groups of people who are lonely or isolated. While the presence of social prescribing codes indicate that conversations are happening between GPs and patients, this analysis is unable to determine whether patients then met a link worker, how long they engaged with the service and whether they subsequently accessed community activities. Our analyses are also limited to primary care records, so do not provide a complete picture of social prescribing among people with SMI in England. Integration between primary care medical records and social prescribing databases such as Joy, Social Rx Connect and Access Elemental (used by link workers to monitor referrals and record their patient appointments) would enable a more comprehensive understanding of the full social prescribing pathway. Further work is also needed to understand the potential impact of social prescribing on health outcomes and on secondary care health service use to establish whether its focus on increasing community support translates into reduced hospitalisations and A&E attendance among people with SMI. Finally, we are unable to determine from the code “social prescribing declined” whether refusals were determined by patients, referrers or link workers and, therefore, whether this represents patient choice or healthcare system level barriers to access. Clarification and simplification of SNOMED codes relating to social prescribing is, therefore, important to improve the quality of future analyses on social prescribing in primary care.

In conclusion, this study suggests that social prescribing consultations have increased among patients with SMI in primary care since the service was officially launched in 2019, and that it is reaching patients with SMI and co-existing health problems, as well as those living in more deprived areas. The analysis revealed demographic disparities in consultations among younger adults and males with SMI as well as social prescribing service refusal among older adults for whom targeted strategies to address inequalities in access to social prescribing may therefore be needed.

## Acknowledgements

The authors would like to thank the project lived experience advisory group for their input into the study.

## Author Contributions

AB and AET formulated the research questions. AB, NL, AET, ARB, ST, DO, DF and FB commented on the study protocol and the design of the study. FB conducted the analysis and wrote the first draft of the methods and results sections of the manuscript. AB edited the methods and results section and drafted the manuscript. AB, NL, ARB and DF commented on the results of the analysis. NL, AET, ARB, ST, DO, DF and FB commented and agreed on the full manuscript.

## Data Availability

Data were obtained from a third party and are not publicly available. All data were obtained from Clinical Practice Research Datalink (CPRD). Data is available from CPRD but is subject to protocol approval and ethical review: https://www.cprd.com/research-applications. Analytical code is available on reasonable request.

## References

1. Halstead S, Cao C, Høgnason Mohr G, Ebdrup BH, Pillinger T, McCutcheon RA, et al. Prevalence of multimorbidity in people with and without severe mental illness: a systematic review and meta-analysis. The Lancet Psychiatry. 2024;11(6):431–42.

2. Office for Health improvement and Disparities. Premature mortality in adults with severe mental illness (SMI) 2023 [Available from: https://www.gov.uk/government/publications/premature-mortality-in-adults-with-severe-mental-illness/premature-mortality-in-adults-with-severe-mental-illness-smi].

3. Nielsen RE, Banner J, Jensen SE. Cardiovascular disease in patients with severe mental illness. Nat Rev Cardiol. 2021;18(2):136–45.

4. Hayes JF, Marston L, Walters K, King MB, Osborn DPJ. Mortality gap for people with bipolar disorder and schizophrenia: UK-based cohort study 2000-2014. Br J Psychiatry. 2017;211(3):175–81.

5. Lawrence EG, Zahid U, Thomson AC, Lau R, Havers L, Biotti F, et al. Psychosocial and Demographic Factors Associated with Physical Multimorbidity in Severe Mental Illness: A Systematic Review. Schizophrenia Bulletin. 2025.

6. Peckham E, Allgar V, Crosland S, Heron P, Johnston G, Newbronner E, et al. Investigating smoking and nicotine dependence among people with severe mental illness during the COVID-19 pandemic: analysis of linked data from a UK Closing the Gap cohort. BJPsych Open. 2021;7(3):e86.

7. Launders N, Hayes JF, Price G, Osborn DPJ. Clustering of physical health multimorbidity in people with severe mental illness: An accumulated prevalence analysis of United Kingdom primary care data. PLOS Medicine. 2022;19(4):e1003976.

8. Li W, Yang Y, An F-R, Zhang L, Ungvari GS, Jackson T, et al. Prevalence of comorbid depression in schizophrenia: A meta-analysis of observational studies. Journal of Affective Disorders. 2020;273:524–31.

9. Henderson C, Kotera Y, Lloyd-Evans B, Jordan G, Gorner M, Salla A, et al. Social inclusion of people with severe mental illness: a review of current practices, evidence and unmet needs, and future directions. World Psychiatry. 2026;25(1):56–82.

10. Naito R, McKee M, Leong D, Bangdiwala S, Rangarajan S, Islam S, et al. Social isolation as a risk factor for all-cause mortality: Systematic review and meta-analysis of cohort studies. PLoS One. 2023;18(1):e0280308.

11. Malogianni E, Alenezi MF, Renwick L. Barriers and facilitators of accessing primary healthcare for patients with severe mental illness: a mixed-methods systematic review using framework synthesis. BMC Psychiatry. 2025;25(1):1131.

12. Lawrence D, Kisely S. Inequalities in healthcare provision for people with severe mental illness. J Psychopharmacol. 2010;24(4 Suppl):61–8.

13. Thomson LJM, Chatterjee HJ. The role of community assets in tackling UK health inequalities through community engagement and partnerships in social prescribing. Frontiers in Public Health. 2026;Volume 14 – 2026.

14. NHS England. Social Prescribing 2023 [Available from: https://www.england.nhs.uk/personalisedcare/social-prescribing/].

15. Wilson A, Noble H, Galway K, Doherty J. Social prescribing for people living with long-term health conditions: a scoping review. Syst Rev. 2025;14(1):114.

16. Oliveira D, Henriques A, Nogueira P, Costa A. Acceptability of a social prescribing program (REDE D+) for self-care and health literacy among people with type 2 diabetes mellitus-a qualitative study. Front Public Health. 2026;14:1762723.

17. Cooper M, Avery L, Scott J, Ashley K, Jordan C, Errington L, et al. Effectiveness and active ingredients of social prescribing interventions targeting mental health: a systematic review. BMJ Open. 2022;12(7):e060214.

18. Reinhardt GY, Vidovic D, Hammerton C. Understanding loneliness: a systematic review of the impact of social prescribing initiatives on loneliness. Perspect Public Health. 2021;141(4):204–13.

19. Barlattani T, Venturiello D, Grassi D, Romano S, Pacitti F. Bridging the heart–mind divide: Psychiatric implementation of the 2025 ESC mental–CVD consensus. European Psychiatry. 2026;69(1):e22.

20. NHS England. Improving the physical health of people living with severe mental illness. Guidance for integrated care systems: NHS England; 2025 [Available from: https://www.england.nhs.uk/long-read/improving-the-physical-health-of-people-living-with-severe-mental-illness/].

21. Leavey G, Watterson R, McAnee G, Shannon S, Boyd K, Lloyd-Evans B, et al. A social prescribing model for tackling the health and social inequalities of people living with severe mental illness: a protocol paper. BMC Public Health. 2025;25(1):3211.

22. Baxter L, Burton A, Fancourt D. Community and cultural engagement for people with lived experience of mental health conditions: what are the barriers and enablers? BMC Psychol. 2022;10(1):71.

23. Aughterson H, Baxter L, Fancourt D. Social prescribing for individuals with mental health problems: a qualitative study of barriers and enablers experienced by general practitioners. BMC Family Practice. 2020;21(1):194.

24. Wilding A, Sutton M, Agboraw E, Munford L, Wilson P. Geographic inequalities in need and provision of social prescribing link workers a retrospective study in primary care. Br J Gen Pract. 2024;74(748):e784–e90.

25. Turk A, Tierney S, Hogan B, Mahtani KR, Pope C. A meta-ethnography of the factors that shape link workers’ experiences of social prescribing. BMC Medicine. 2024;22(1):280.

26. Hazeldine E, Gowan G, Wigglesworth R, Pollard J, Asthana S, Husk K. Link worker perspectives of early implementation of social prescribing: A ‘Researcher-in-Residence’ study. Health Soc Care Community. 2021;29(6):1844–51.

27. Wolf A, Dedman D, Campbell J, Booth H, Lunn D, Chapman J, et al. Data resource profile: Clinical Practice Research Datalink (CPRD) Aurum. Int J Epidemiol. 2019;48(6):1740–g.

28. Mahadevan P, Harley M, Fordyce S, Hodgson S, Ghosh R, Myles P, et al. Completeness and representativeness of small area socioeconomic data linked with the UK Clinical Practice Research Datalink (CPRD). J Epidemiol Community Health. 2022;76(10):880–6.

29. NHS England. The NHS Long Term Plan 2019 [Available from: https://webarchive.nationalarchives.gov.uk/ukgwa/20230418155402/https:/www.longtermplan.nhs.uk/publication/nhs-long-term-plan/].

30. Bu F, Burton A, Launders N, Taylor AE, Richards-Belle A, Tierney S, et al. National roll-out of social prescribing in England’s primary care system: a longitudinal observational study using Clinical Practice Research Datalink data. The Lancet Public Health. 2025;10(11):e903–e11.

31. Wilding A, Agboraw E, Munford L, Sutton M, Mercer SW, Salisbury C, et al. Impact of the rollout of the national social prescribing link worker programme on population outcomes: evidence from a repeated cross-sectional survey. British Journal of General Practice. 2025;75(761):e880–e8.

32. Dayson C, Painter J, Bennett E. Social prescribing for patients of secondary mental health services: emotional, psychological and social well-being outcomes. Journal of Public Mental Health. 2020;19(4):271–9.

33. Khan K, Al-Izzi R, Montasem A, Gordon C, Brown H, Goldthorpe J. The feasibility of identifying health inequalities in social prescribing referrals and declines using primary care patient records. NIHR Open Res. 2023;3:.

34. Hardoon S, Hayes JF, Blackburn R, Petersen I, Walters K, Nazareth I, et al. Recording of Severe Mental Illness in United Kingdom Primary Care, 2000–2010. PLOS ONE. 2013;8(12):e82365.

35. Aleman A, Kahn RS, Selten J-P. Sex Differences in the Risk of Schizophrenia: Evidence From Meta-analysis. Archives of General Psychiatry. 2003;60(6):565–71.

36. Poole R, Huxley P. Social prescribing: an inadequate response to the degradation of social care in mental health. BJPsych Bulletin. 2024;48(1):30–3.

37. Office for Health improvement and Disparities. Estimated prevalence of severe mental illness in England: report UK: Department of Health and Social Care; 2026 [Available from: https://www.gov.uk/government/statistics/adult-mental-health-and-wellbeing-profile-april-2026-update/estimated-prevalence-of-severe-mental-illness-in-england-report.

38. Lau JYF, Priebe S, Morgan C. Social Health and Serious Mental Illness-A Step Forward? JAMA Psychiatry. 2025;82(3):213–4.

39. Sadio R, Henriques A, Nogueira P, Costa A. Barriers and Facilitators to Older Adults’ Engagement in Social Prescribing: A Qualitative Study Using Focus Groups. J Clin Med. 2025;14(13).

40. Fortuna KL, Lohman MC, Batsis JA, DiNapoli EA, DiMilia PR, Bruce ML, et al. Patient experience with healthcare services among older adults with serious mental illness compared to the general older population. Int J Psychiatry Med. 2017;52(4-6):381–98.

