## Supplementary material for "Social prescribing for patients with severe mental illnesses: an English primary care longitudinal observational study": Tables and figures

Table 1. Patient characteristics in each study cohort

|  |  | Cohort 1 (SMI diagnosis before 2019)  (n=112,334) | | Cohort 2 (SMI diagnosis up to and including 2023)  (n=140,514) | |
| --- | --- | --- | --- | --- | --- |
|  |  | Freq. | % | Freq. | % |
| Age groups: | Under 30 | 12,535 | 11.2% | 21,964 | 15.6% |
|  | 30-39 | 21,396 | 19.0% | 27,847 | 19.8% |
|  | 40-49 | 24,919 | 22.2% | 29,527 | 21.0% |
|  | 50-59 | 25,752 | 22.9% | 29,156 | 20.7% |
|  | 60-69 | 15,409 | 13.7% | 17,411 | 12.4% |
|  | 70+ | 12,323 | 11.0% | 14,609 | 10.4% |
| Sex: | Males | 59,141 | 52.6% | 72,976 | 51.9% |
|  | Females | 53,193 | 47.4% | 67,538 | 48.1% |
| Ethnicity: | Ethnic minority | 24,289 | 23.1% | 31,533 | 24.0% |
|  | White | 80,978 | 76.9% | 100,101 | 76.0% |
| IMD quintiles: | 1 (least deprived) | 11,083 | 12.1% | 13,801 | 12.1% |
|  | 2 | 13,484 | 14.7% | 16,712 | 14.7% |
|  | 3 | 16,603 | 18.1% | 20,598 | 18.1% |
|  | 4 | 22,939 | 25.1% | 28,681 | 25.2% |
|  | 5 (most deprived) | 27,437 | 30.0% | 34,097 | 29.9% |
| Urbanicity: | Urban | 82,311 | 89.9% | 102,544 | 90.0% |
|  | Rural | 9,235 | 10.1% | 11,345 | 10.0% |
| BMI: | Under/normal weight | 32,734 | 31.1% | 42,402 | 33.6% |
|  | Overweight | 33,582 | 31.9% | 39,446 | 31.3% |
|  | Obese | 39,104 | 37.1% | 44,372 | 35.2% |
| Diabetes: | Yes | 14,814 | 13.2% | 16,338 | 11.6% |
|  | No | 97,520 | 86.8% | 124,176 | 88.4% |
| Hypertension: | Yes | 19,785 | 17.6% | 22,737 | 16.2% |
|  | No | 92,549 | 82.4% | 117,777 | 83.8% |
| Depression: | Yes | 51,331 | 45.7% | 62,491 | 44.5% |
|  | No | 61,003 | 54.3% | 78,023 | 55.5% |

Table 2. Results from the fixed effects models treating SMI as time-varying (n=140,514 N=606,622)

| Variable | OR | 95% CI | P |
| --- | --- | --- | --- |
| SMI (yes vs no) | 1.39 | [1.26, 1.53] | <0.001 |
| Year: 2020 (vs 2019) | 5.43 | [4.54, 6.49] | <0.001 |
| Year: 2021 (vs 2019) | 7.84 | [6.60, 9.31] | <0.001 |
| Year: 2022 (vs 2019) | 13.2 | [11.02, 15.88] | <0.001 |
| Year: 2023 (vs 2019) | 15.2 | [12.62, 18.30] | <0.001 |

Notes: n denotes the number of patients; N denotes the number of patient-year observations

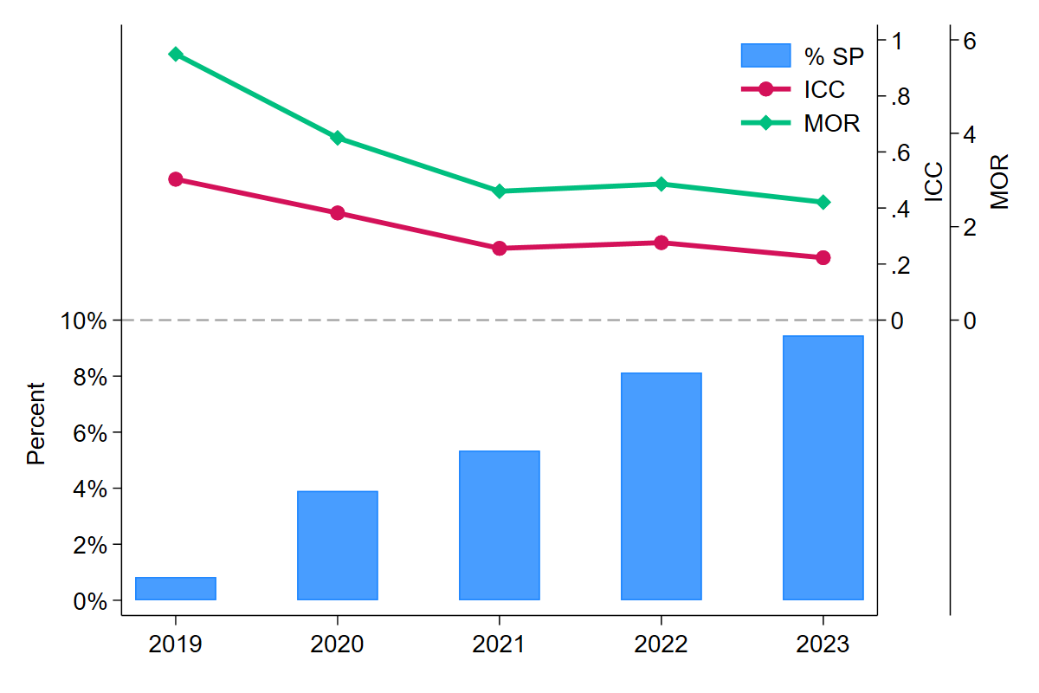

Figure 1. Percentage of SMI patients with SP consultation Intraclass correlation coefficients (ICC) from unconditional multilevel logistic models by year

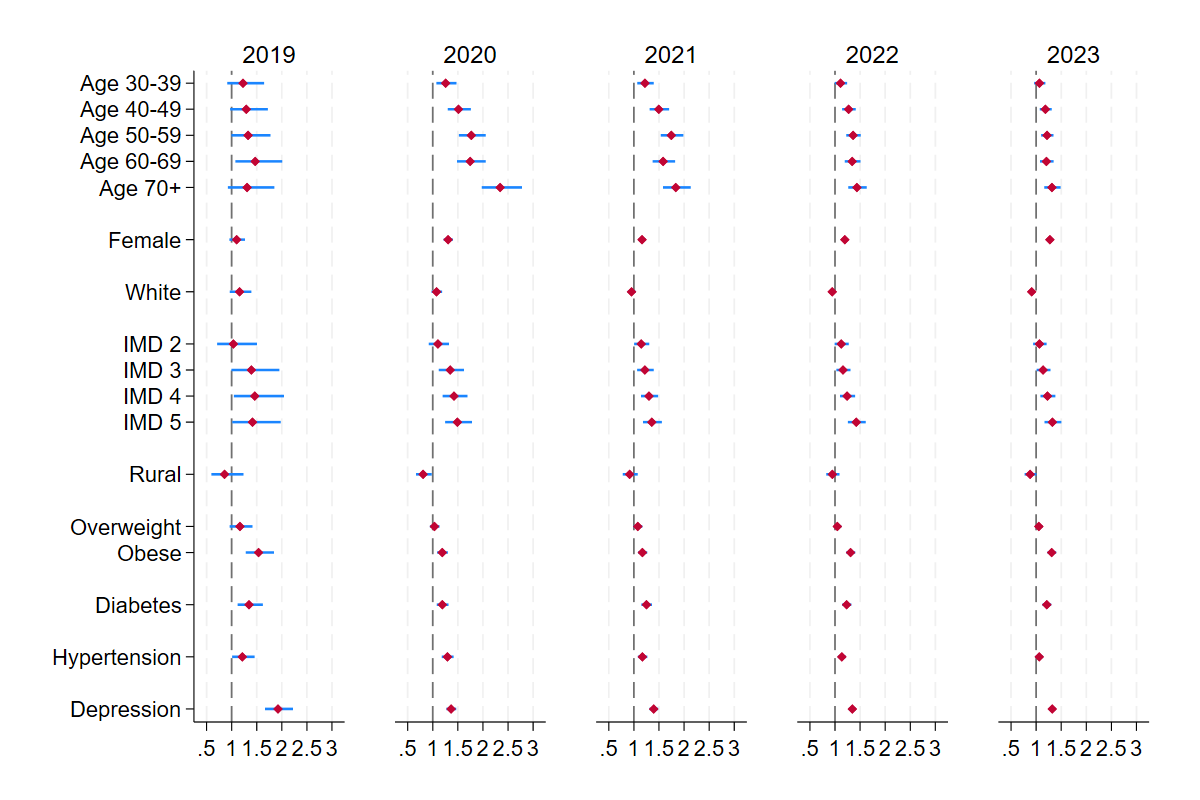

(a) **SP Consultations**

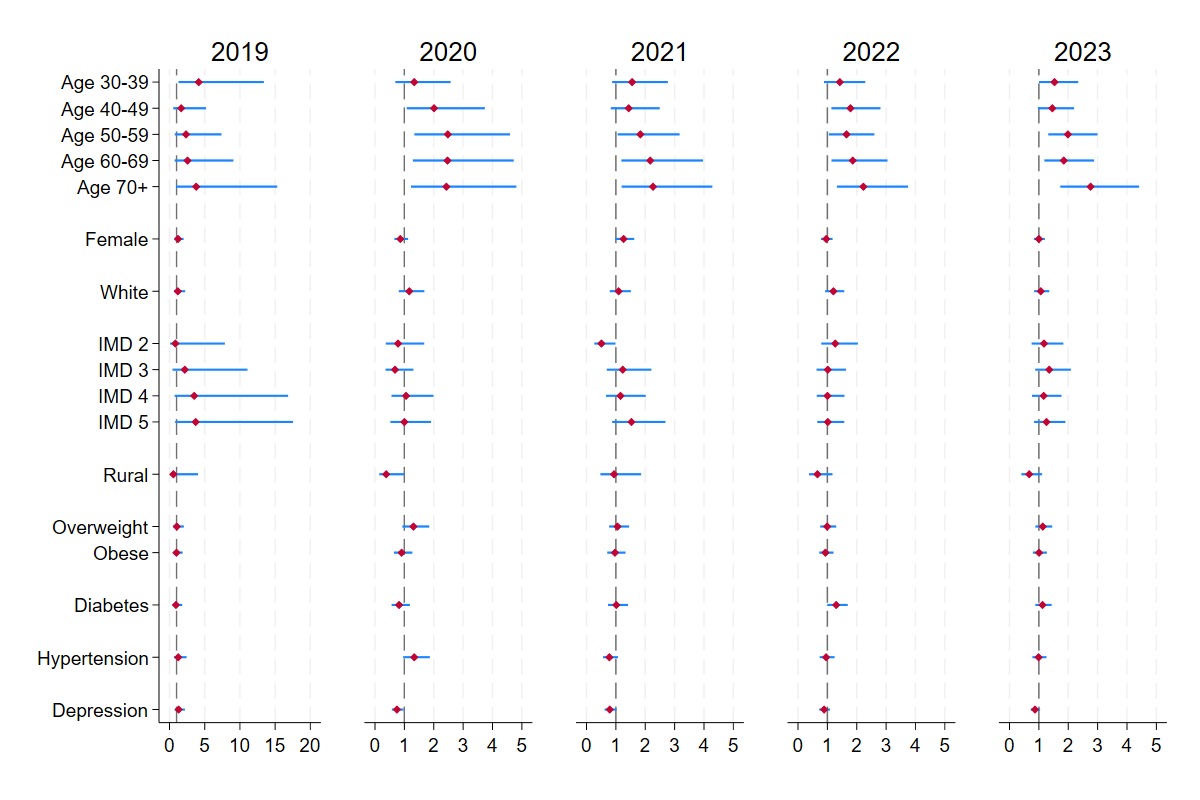

**(b) SP Service Refusals**

Figure 2. Estimated odds ratio and 95% confidence intervals from multilevel logistic regression models stratified by year for (a) SP consultations and (b) SP service refusals
