## Supplementary Materials 1 for "Social prescribing for patients with severe mental illnesses: an English primary care longitudinal observational study"

Table S1. SNOMED codes related to social prescribing

| **MedCodeId** | **SNOMED CT terms** |
| --- | --- |
| 2252091000000114 | Referral to social prescribing service |
| 2252011000000117 | Social prescribing offered |
| 14308041000006115 | Seen by social prescribing link worker |
| 2717001000000119 | Signposting to social prescribing service |
| 1969731000006116 | Review of social prescribing plan |
| 2252051000000118 | Social prescribing declined |
| 14233901000006110 | Social prescribing case closed |
| 1969741000006114 | Social prescribing plan completed |
| 1148481000000117 | Social prescribing for mental health |
| 11932261000006119 | Not suitable for social prescribing |
| 1969751000006111 | Referral to social prescribing service from other agency |

Table S2. Percentages of missing data in each cohort

|  | Cohort1 | Cohort2 |
| --- | --- | --- |
| Age groups | 0.0% | 0.0% |
| Sex | 0.0% | 0.0% |
| Ethnicity | 6.3% | 6.3% |
| IMD | 18.5% | 18.9% |
| Urbanicity | 18.5% | 18.9% |
| Obesity | 6.2% | 10.2% |
| Diabetes | 0.0% | 0.0% |
| Hypertension | 0.0% | 0.0% |
| Depression | 0.0% | 0.0% |

Table S3 Results from multilevel logistic regression models on SP consultations by year

|  | Model 2019 | | Model 2020 | | Model 2021 | | Model 2022 | | Model 2023 | |
| --- | --- | --- | --- | --- | --- | --- | --- | --- | --- | --- |
| Age 30-39 (vs <30) | 1.23 | [0.91,1.65] | 1.26 | [1.07,1.47] | 1.22 | [1.07,1.40] | 1.11 | [0.99,1.24] | 1.07 | [0.96,1.18] |
| Age 40-49 (vs <30) | 1.29 | [0.97,1.72] | 1.51 | [1.30,1.76] | 1.50 | [1.32,1.70] | 1.27 | [1.14,1.41] | 1.18 | [1.07,1.31] |
| Age 50-59 (vs <30) | 1.33 | [0.99,1.77] | 1.77 | [1.52,2.06] | 1.75 | [1.54,1.99] | 1.36 | [1.22,1.51] | 1.22 | [1.10,1.35] |
| Age 60-69 (vs <30) | 1.47 | [1.07,2.01] | 1.75 | [1.48,2.06] | 1.58 | [1.37,1.82] | 1.34 | [1.19,1.51] | 1.21 | [1.08,1.35] |
| Age 70+ (vs <30) | 1.31 | [0.92,1.85] | 2.34 | [1.98,2.78] | 1.84 | [1.58,2.13] | 1.44 | [1.26,1.63] | 1.31 | [1.16,1.49] |
| Female (vs male) | 1.10 | [0.95,1.27] | 1.31 | [1.22,1.40] | 1.16 | [1.09,1.24] | 1.20 | [1.13,1.26] | 1.27 | [1.21,1.34] |
| White (vs other) | 1.16 | [0.96,1.39] | 1.07 | [0.97,1.19] | 0.95 | [0.88,1.03] | 0.95 | [0.88,1.01] | 0.91 | [0.85,0.98] |
| IMD 2 (vs 1) | 1.03 | [0.71,1.51] | 1.10 | [0.92,1.32] | 1.15 | [1.01,1.31] | 1.12 | [0.99,1.27] | 1.07 | [0.94,1.21] |
| IMD 3 (vs 1) | 1.39 | [0.99,1.95] | 1.35 | [1.12,1.62] | 1.22 | [1.06,1.39] | 1.16 | [1.03,1.31] | 1.14 | [1.01,1.29] |
| IMD 4 (vs 1) | 1.46 | [1.05,2.04] | 1.42 | [1.20,1.69] | 1.30 | [1.14,1.48] | 1.24 | [1.10,1.40] | 1.23 | [1.09,1.38] |
| IMD 5 (vs 1) | 1.42 | [1.01,1.98] | 1.49 | [1.25,1.78] | 1.36 | [1.18,1.56] | 1.42 | [1.26,1.61] | 1.32 | [1.17,1.50] |
| Rural (vs urban) | 0.86 | [0.60,1.24] | 0.81 | [0.67,0.98] | 0.92 | [0.78,1.08] | 0.95 | [0.82,1.09] | 0.88 | [0.77,1.01] |
| Overweight (vs other) | 1.17 | [0.96,1.42] | 1.03 | [0.94,1.13] | 1.08 | [0.99,1.17] | 1.05 | [0.97,1.12] | 1.05 | [0.98,1.13] |
| Obese (vs other) | 1.54 | [1.28,1.84] | 1.19 | [1.09,1.30] | 1.17 | [1.08,1.27] | 1.31 | [1.22,1.40] | 1.31 | [1.23,1.40] |
| Diabetes (yes vs no) | 1.35 | [1.12,1.62] | 1.19 | [1.08,1.31] | 1.25 | [1.15,1.36] | 1.23 | [1.14,1.33] | 1.21 | [1.13,1.30] |
| Hypertension (yes vs no) | 1.21 | [1.01,1.46] | 1.29 | [1.18,1.42] | 1.17 | [1.08,1.27] | 1.14 | [1.06,1.22] | 1.06 | [0.99,1.14] |
| Depression (yes vs no) | 1.92 | [1.66,2.22] | 1.37 | [1.27,1.47] | 1.39 | [1.31,1.48] | 1.35 | [1.27,1.42] | 1.32 | [1.25,1.39] |
| Var (_cons) | 32.04 | [15.93,64.45] | 8.15 | [6.29,10.56] | 3.24 | [2.82,3.71] | 3.64 | [3.18,4.16] | 2.59 | [2.34,2.87] |
| n (patient) | 108,582 | | 101,175 | | 96,044 | | 90,271 | | 83,086 | |
| N (practice) | 1,770 | | 1,734 | | 1,700 | | 1,677 | | 1,615 | |
| % of having SP consultations | 0.8% | | 3.9% | | 5.3% | | 8.1% | | 9.5% | |

Table S4 Results from multilevel logistic regression models on SP service refusal by year

|  | Model 2019 | | Model 2020 | | Model 2021 | | Model 2022 | | Model 2023 | |
| --- | --- | --- | --- | --- | --- | --- | --- | --- | --- | --- |
| Age 30-39 (vs <30) | 4.15 | [1.28,13.41] | 1.34 | [0.69,2.57] | 1.55 | [0.87,2.77] | 1.42 | [0.89,2.29] | 1.53 | [1.00,2.34] |
| Age 40-49 (vs <30) | 1.66 | [0.53,5.20] | 2.01 | [1.08,3.74] | 1.44 | [0.83,2.50] | 1.79 | [1.14,2.81] | 1.46 | [0.97,2.20] |
| Age 50-59 (vs <30) | 2.34 | [0.74,7.38] | 2.48 | [1.34,4.60] | 1.83 | [1.06,3.17] | 1.65 | [1.05,2.60] | 1.99 | [1.32,3.00] |
| Age 60-69 (vs <30) | 2.56 | [0.72,9.06] | 2.47 | [1.29,4.73] | 2.17 | [1.19,3.97] | 1.86 | [1.14,3.05] | 1.85 | [1.19,2.88] |
| Age 70+ (vs <30) | 3.79 | [0.94,15.31] | 2.43 | [1.23,4.82] | 2.27 | [1.20,4.29] | 2.23 | [1.32,3.75] | 2.76 | [1.73,4.42] |
| Female (vs male) | 1.21 | [0.73,2.01] | 0.86 | [0.66,1.13] | 1.26 | [0.98,1.63] | 0.96 | [0.78,1.18] | 1.00 | [0.83,1.20] |
| White (vs other) | 1.18 | [0.63,2.21] | 1.17 | [0.81,1.68] | 1.09 | [0.79,1.51] | 1.21 | [0.92,1.57] | 1.07 | [0.84,1.35] |
| IMD 2 (vs 1) | 0.83 | [0.09,7.86] | 0.79 | [0.37,1.68] | 0.51 | [0.26,0.99] | 1.27 | [0.79,2.04] | 1.18 | [0.75,1.84] |
| IMD 3 (vs 1) | 2.17 | [0.42,11.08] | 0.68 | [0.36,1.31] | 1.23 | [0.69,2.21] | 1.02 | [0.63,1.64] | 1.35 | [0.88,2.09] |
| IMD 4 (vs 1) | 3.51 | [0.73,16.85] | 1.06 | [0.56,1.99] | 1.16 | [0.66,2.02] | 1.00 | [0.64,1.58] | 1.16 | [0.77,1.77] |
| IMD 5 (vs 1) | 3.71 | [0.79,17.55] | 1.00 | [0.53,1.90] | 1.53 | [0.87,2.69] | 1.02 | [0.65,1.58] | 1.26 | [0.83,1.90] |
| Rural (vs urban) | 0.55 | [0.07,4.04] | 0.38 | [0.14,1.01] | 0.94 | [0.47,1.86] | 0.66 | [0.37,1.17] | 0.67 | [0.40,1.11] |
| Overweight (vs other) | 1.03 | [0.53,2.03] | 1.31 | [0.93,1.85] | 1.06 | [0.77,1.45] | 0.99 | [0.76,1.30] | 1.13 | [0.88,1.45] |
| Obese (vs other) | 0.98 | [0.52,1.86] | 0.91 | [0.64,1.27] | 0.97 | [0.71,1.33] | 0.93 | [0.72,1.21] | 1.01 | [0.79,1.27] |
| Diabetes (yes vs no) | 0.91 | [0.46,1.81] | 0.82 | [0.57,1.19] | 1.02 | [0.73,1.41] | 1.30 | [1.00,1.69] | 1.13 | [0.88,1.43] |
| Hypertension (yes vs no) | 1.24 | [0.63,2.43] | 1.34 | [0.96,1.87] | 0.78 | [0.56,1.08] | 0.96 | [0.73,1.25] | 0.99 | [0.78,1.26] |
| Depression (yes vs no) | 1.29 | [0.76,2.18] | 0.75 | [0.57,0.98] | 0.79 | [0.62,1.02] | 0.89 | [0.73,1.09] | 0.87 | [0.72,1.04] |
| Var (_cons) | 528.64 | [2.76,101258.17] | 429.49 | [59.89,3079.84] | 42.44 | [13.14,137.06] | 44.72 | [17.21,116.22] | 18.03 | [9.49,34.22] |
| n (patient) | 899 | | 3,941 | | 5,132 | | 7,342 | | 7,851 | |
| N (practice) | 58 | | 279 | | 418 | | 553 | | 680 | |
| % declined | 16.1% | | 14.0% | | 9.4% | | 10.1% | | 10.8% | |

Table S5 Panel descriptive statistics of SMI and SP (2019-2023)

|  |  | Overall | | Between | | Within |
| --- | --- | --- | --- | --- | --- | --- |
|  |  | Freq. | Percent | Freq. | Percent | Percent |
| SMI | No | 47134 | 7.8% | 20635 | 14.7% | 49.0% |
|  | Yes | 559488 | 92.2% | 140514 | 100.0% | 92.8% |
|  | Total | 606622 | 100.0% | 161149 | 114.7% | 87.2% |
| SP | No | 574665 | 94.7% | 140408 | 99.9% | 95.3% |
|  | Yes | 31957 | 5.3% | 23885 | 17.0% | 28.0% |
|  | Total | 606622 | 100.0% | 164293 | 116.9% | 85.5% |

Notes: Overall statistics describe the distribution across all person-year observations. Between-person statistics describes the number and percentage of individuals observed in each category at least once across years. Within-person percentages indicate the average proportion of observed person-years spent in each category among individuals who were observed in that category at least once.

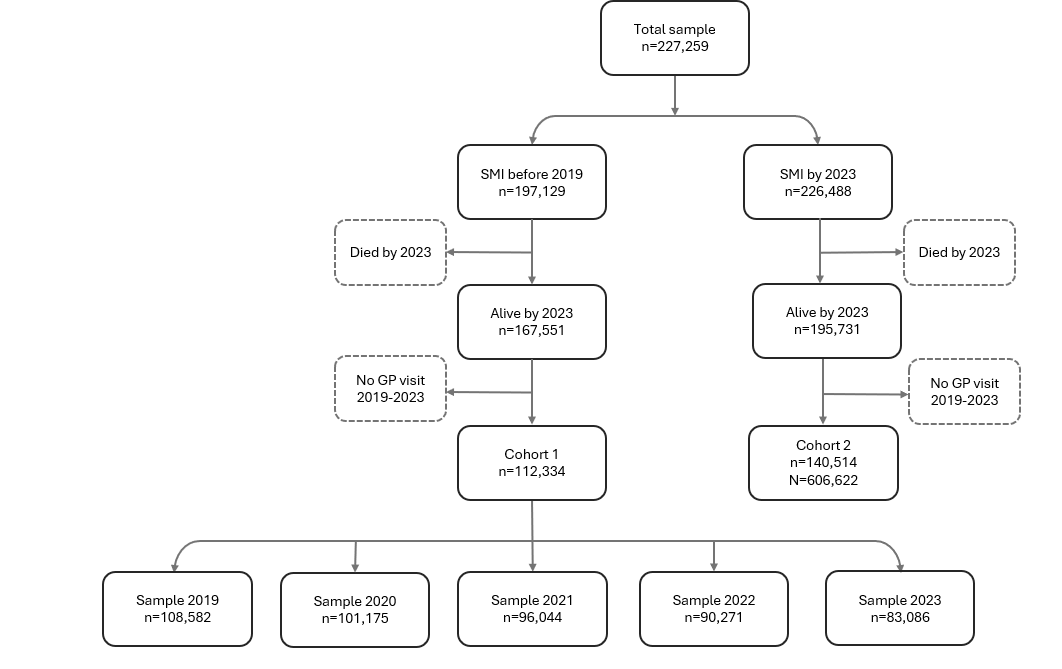
Figure S1. Sample selection diagram
